# Detection of Circulating Tumor-specific DNA Methylation Markers in the Blood of Patients with Pituitary Tumors

**DOI:** 10.1101/2020.05.29.20116202

**Authors:** Michael Wells, Karam P. Asmaro, Thais S. Sabedot, Tathiane M. Malta, Maritza S. Mosella, Kevin Nelson, James Snyder, Ana deCarvalho, Abir Mukherjee, Dhananjay Chitale, Adam Robin, Mark Rosenblum, Thomas Mikkelsen, Laila M. Poisson, Ian Y. Lee, Tobias Walbert, Arti Bhan, Steven Kalkanis, Jack Rock, Houtan Noushmehr, Ana Valeria Castro

## Abstract

Genome-wide DNA methylation aberrations are pervasive and associated with clinicopathological features across pituitary tumors (PT) subtypes. The feasibility to detect CpG methylation abnormalities in circulating cell-free DNA (cfDNA) has been reported in central nervous system tumors other than PT. Here, we aimed to profile and identify methylome-based signatures in the serum of patients harboring PT (n =13). Our analysis indicated that serum cfDNA methylome from patients with PT are distinct from the counterparts in patients with other tumors (gliomas, meningiomas, colorectal carcinomas, n =134) and nontumor conditions (n = 4). Furthermore, the serum methylome patterns across PT was associated with functional status and adenohypophyseal cell lineage PT subtypes, recapitulating epigenetic features reported in PT-tissue. A machine learning algorithm using serum PT-specific signatures generated a score that distinguished PT from non-PT conditions with 100% accuracy in our validation set. These preliminary results underpin the potential clinical application of a liquid biopsy-based DNA methylation profiling as a noninvasive approach to identify clinically relevant epigenetic markers that can be used in the management of PT.

## Introduction

Liquid biopsy (LB) is a method used to detect molecular elements (e.g., DNA, RNA, etc.) shed by tumors in biofluids (blood, cerebrospinal fluid etc). Circulating cell-free DNA (cfDNA), specifically the tumor DNA fraction (ctDNA), is thought to originate from cellular death (apoptosis and necrosis) or secretion from live cells, especially from proliferative tissues or tumors ^1–5^. Blood-based LB has emerged as a reliable and a minimally invasive approach to identify clinically relevant molecular biomarkers from several tumor origins, including from central nervous system (CNS)neoplasms ^5–8^.

In contrast to CNS tumors that are shielded by the blood-brain barrier, the pituitary gland presents an anatomical structure that facilitates the spillage of tumor cellular material into the bloodstream, i.e. a fenestrated pituitary portal system and/or an access to the cavernous system. This structural advantage creates an opportunity to profile tumor-specific molecular features of material released from these tumors potentially suitable for clinicopathological application ^9,10^. Indeed, the feasibility to detect and sequence somatic gene variants in ctDNA has recently been reported in PT ^1^; however, the detection sensitivity of this approach was low in these tumors ^1^. The paucity of genetic alterations in the pathogenesis of PT such as recurrent somatic mutations may have contributed to these results^11–15^. In contrast, genome-wide methylation abnormalities detected in the tissue are knowingly pervasive across PT subtypes ^13,16–23^.Additionally,DNA methylome patterns are tissue- and tumor-specific providing an opportunity to predict the tissue of origin of the tumor through DNA methylation profiling ^5,20,24,25^. In fact, many studies showed that specific methylome patterns detected in the tissue distinguished PT from other CNS tumors and defined discrete methylation subtypes among different CNS tumors ^20,26,27^. Additionally, methylation markers presented diagnostic, prognostic and predictive applications in CNS tumors ^20,26,27^. The feasibility of detecting these tissue- or tumor-specific methylation signatures using a liquid biopsy approach is an emerging field that has not been reported in PT to date.

In this study, we profiled the serum cfDNA methylome derived from patients with PT or other tumors and nontumor conditions. We identify unique methylation signatures in the serum associated with clinicopathological features specific to PT. This proof-of concept study paves the way for the potential clinical application of a liquid biopsy as a noninvasive approach to identify and assess relevant epigenetic markers that may be useful in the management of patients with PT.

## Results

### Characterization of pituitary cell-free DNA methylome cfDNA quantification

Total extracted serum cfDNA quantity, normalized to the genomic size (ng/ml, see Methods), were not significantly different from controls (mean±SD, 59.3±134.2 vs 5±5.0 ng/ml, respectively; p=.14) or in relation to functional or invasion status in PT **(Supplemental Figure 1A)**.

### Deconvolution

The deconvolution of the serum cfDNA methylome showed that patients harboring PT had higher proportion of bulk pituitary gland signatures compared to the control serum and other CNS conditions (3%higher, p =.05)**(Supplemental Figure 1B; Supplemental Table 2)**.

### Methylome analysis

The genome-wide mean methylation landscape of the serum cfDNA from patients with PT and non-PT conditions (gliomas, meningiomas, colorectal carcinomas and nontumor conditions) showed that PT segregated into a hypomethylated and a hypermethylated cluster; the latter, shared similar CpG methylation degree with the serum methylome from patients with glioma, meningioma, colorectal cancer and nontumor controls **(Figure 1A and 1B)**.

**Figure 1.**
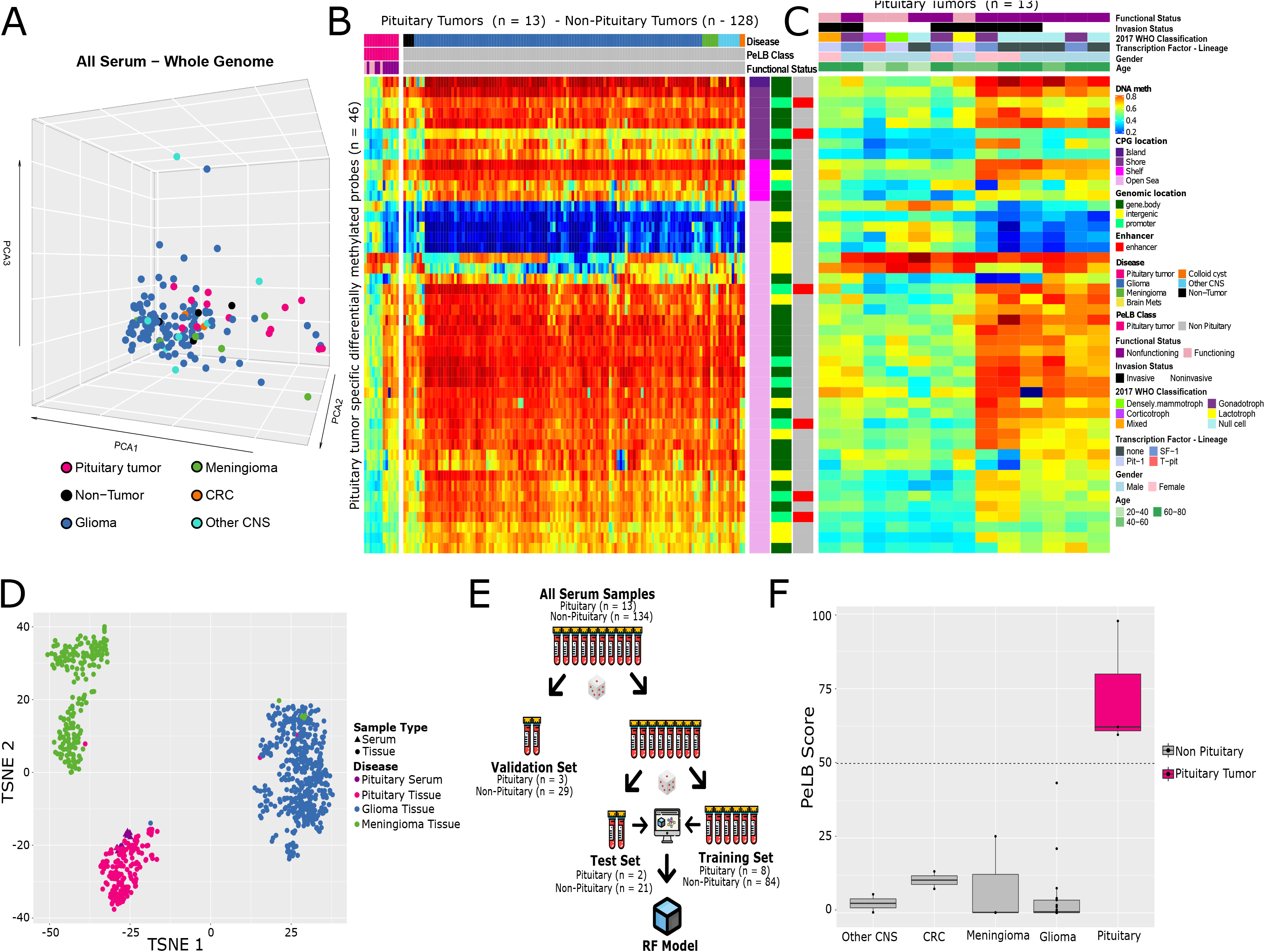
Genome-wide DNA methylation profile of pituitary serum cfDNA. Identification of tissue-specific probes that distinguish pituitary serum from non-pituitary serum specimens (<u>P</u>ituitary-tumor specific <u>E</u>pigenetic <u>L</u>iquid-<u>b</u>iopsy or PeLB probes).(A) PCA of the mean methylation of pituitary tumors and non-pituitary tumors (glioma, meningioma and colorectal carcinoma) and controls (nontumor conditions). (B)Heatmap depicting the methylation levels of PeLB probes across the entire serum cohort (n=107); (C) Heatmap highlights the methylation levels of PeLB probes across pituitary samples and overlapping clinicopathological annotations such as functional and invasion status, transcription factor-related adenohypophyseal cell lineage. (D) The t-Distributed Stochastic Neighbor Embedding (t-SNE) plot depictsthe overlap of PeLB probes with the methylome of serum specimens from patients harboring pituitary tumors and of the tissue methylome from PT and other CNS tumors (glioma, meningioma). (E) Workflow of sample partitioning of serum cohort for training, testing, and validation used in the random forest analysis to distinguish pituitary tumors from non-pituitary tumors (colorectal carcinoma, glioma, meningiomas). Each test tube represents 10% of the samples. (F) Boxplot of the PeLB score results from the validation set. The dottedline at 50% represents the cutoff used for classification into pituitary tumors and non-pituitary samples.

Conducting a supervised analysis between PT and non-PT serum specimens and selecting probes that shared similarities with the matching PT tissue **(Supplemental Figure 1D, left)**, we identified 46 differentially methylated probes (DMP), namely Pituitary Tumors-specific Epigenetic-Liquid Biopsy (PeLB) probes, that significantly distinguished both groups **(Figure 1B)**and distinguished two subgroups across PT (hyper and hypomethylated) (Figure 1C),

The two methylation clusters were associated with distinct clinicopathological status, i.e. the hypermethylated cluster was predominantly composed of nonfunctioning, mainly encompassed by SF1 lineage and null cell tumors and the hypomethylated with functioning PT mostly comprised of Pit1-lineage tumors (Figure 1C). As an exception, one lactotroph adenoma/Pit1-lineage segregated with nonfunctioning PT, despite being clinically classified as functioning, and a functioning Tpit1-lineage tumor clustered with nonfunctioning tumors. These results recapitulate the findings in their matching tissue (Supplemental Figure 1E).

### cfDNA methylome from patients with PT pituitary-specific epigenetic signatures distinct from other pathological conditions

Overlapped with tumor tissues, PeLB probes clustered with PT tissue and significantly separated PT from other CNS tumor-tissue, confirming PeLB-specificity to PT in an independent cohort (**Figure 1D)**. Taking this feature into account, we developed and cross-validated a score derived from a machine learning (ML) model (repeated 5000 times), namely the PeLB score, to predict whether a serum specimen originates from a patient with PT or a non-PT condition **(Figures 1E–F)**. Pituitary-derived serum methylome samples carried the highest values of PeLB score (71–99%), whereas the serum of non-PT tumors carried the lowest values (0–45%) **(Figure F)**. The evaluation of the model in the validation sample set showed that the model performed with an accuracy of 100%, taking into account a 50% PeLB cutoff.

We also defined serum-based methylation signatures (n=70) accounting for the functional/lineage status of PT (nonfunctioning vs functioning PT) (p <.01, differential mean methylation >.2, FDR <.26), we named functioning-PeLB (Func-PeLB)**(Figures 2A, Supplemental Figure 1D, right)**. Harnessing the methylome from matching tissue and publicly available data reporting on the functional status of PT, we observed that a subset of the Func-PeLB probes (overlapped with the 450K platform, used to profile the tissue-methylome of those samples) (n=22 probes) **(Supplemental Figure 1D, right)**, also discriminated the two functional groups at the tissue and respective serum levels (Figure 2B–D)

The CpG probes that distinguished the methylation clusters either in tissue (n= 5000) or serum (6000) were most frequently located in open sea regions (67% and 61%, respectively) and gene bodies (61 and 55%, respectively) **(Figure 1B**,**Supplemental Table 2)**.

## Discussion

Methylome-derived signatures define molecular subtypes that are useful for the diagnosis and prognostication across many tumors ^13,16–18,20,21,26–29^. Additionally, genome-wide DNA methylation patterns are cell-specific either in healthy or tumor specimens^5,18,20,24,25,30–32^. The ability to detect methylation signatures and tumor-specific abnormalities by the profiling of circulating cell-free DNA (cfDNA) in biofluids (liquid biopsy), such as blood, has been useful for the early detection and surveillance of malignant neoplasms ^5,33–35^. In relation to CNS tumors, our group has recently reported on the feasibility to identify methylation-based markers in serum-derived cfDNA for the diagnosis and prognostication of gliomas and meningiomas ^36^. Herein, we show that, similar to malignant and other CNS tumors, PT releases tumor-related information in the blood that allows the identification of clinically relevant methylation signatures specific to patients with PT, namely PeLB probes **(Figure 1A–B, Supplemental Figure 1D)**.Capitalizing on the specificity of these probes, we used a machine learning approach to generate the PeLB score (Figure 1D–E) to predict the presence of a pituitary tumor using liquid biopsy. We showed that PeLB score performed with a 100% accuracy to predict that serum was derived from patients with PT in our validation cohort (**Figure 1F)**. These results remain to be confirmed in an independent cohort of PT-derived, currently unavailable.

In addition, distinct serum DNA methylation landscape, specifically PeLB probes, defined two methylation groups that recapitulated the clinicopathological findings displayed in their matching tissue as reported in other studies ^13,16–18,20,21^**(Figure 1C–D**,**Supplemental Figure 1E)**. These serum-derived clusters showed that the hypermethylated group was enriched by nonfunctioning PT mainly originated from SF1 and Tpit cell lineages and the hypomethylated set mainly composed of functioning PT mostly originated from Pit-1 cell lineages **(Figure 1C, Supplemental Figure 1D**). We narrowed down to a subset of PeLB probes (Func-PeLB) that preserved the distinction between both clusters in tumor-tissue specimens as well **(Figure 2C, Supplemental Figure 1D**). Altogether, these results suggest that PT releases DNA methylation markers in the serum that reflect clinicopathological features such as functional status and adenohypophyseal lineage of these tumors. Confirmation of these findings in a larger and more comprehensive cohort lay the groundwork to the application of PeLB probes as an objective approach to classify PT according to cell-lineage as recommended by the 2017 WHO^37^.

**Figure 2:**
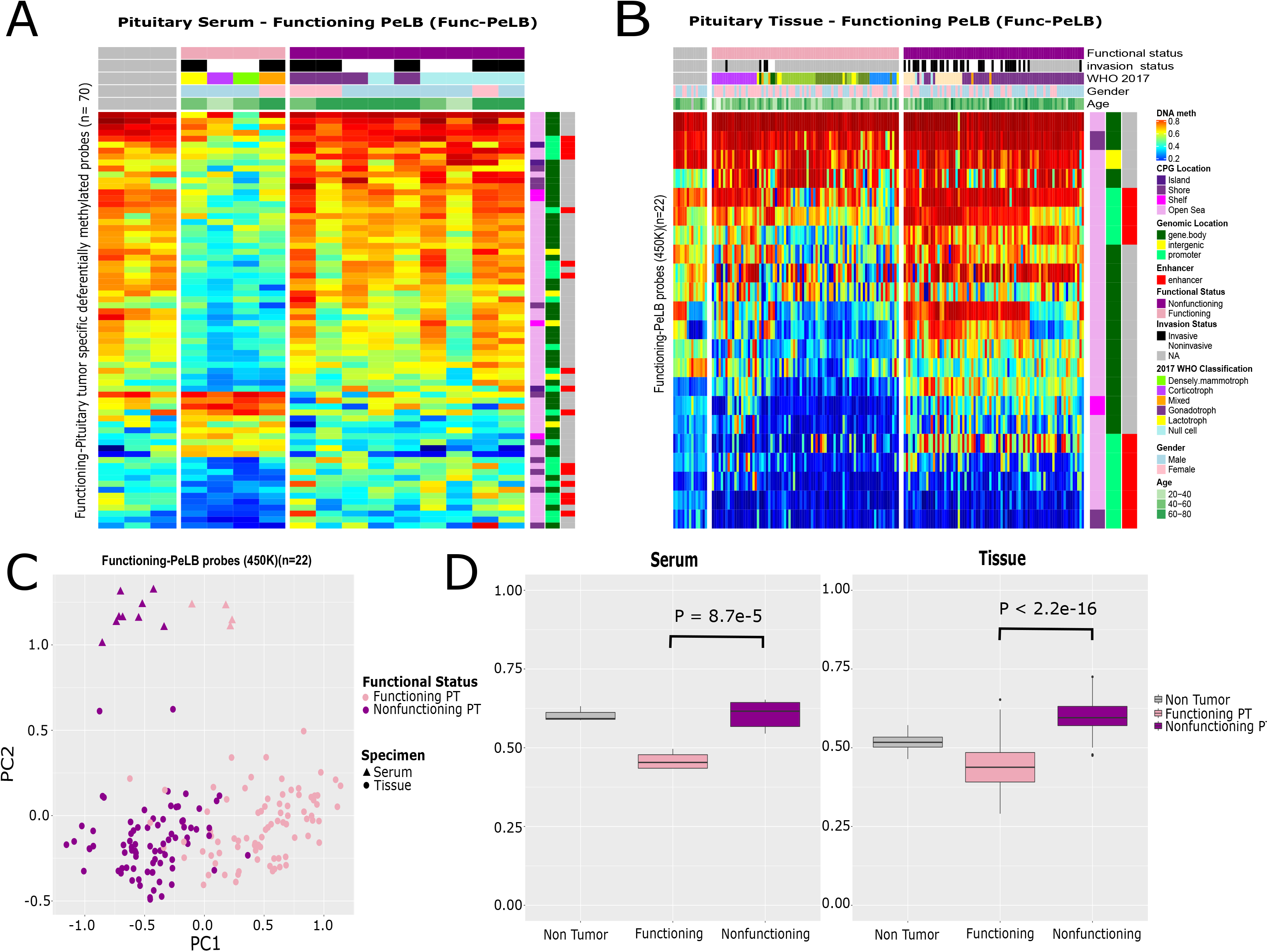
Supervised analysis to identify tissue-specific probes that distinguish serum originated from functioning pituitary tumor from those from nonfunctioning PT (Functioning <u>P</u>ituitary-tumor specific <u>E</u>pigenetic <u>L</u>iquid-<u>b</u>iopsy-PeLB or Func-PeLB). (A) Heatmap displays the methylation levels of the 70 Func-PeLB differentially methylated probes (DMP) in the serum of nontumor and tumor pituitary specimens. (B) Heatmap displays the 22 Func-PeLB porbes that overlap with the 450K array in nontumor and pituitary tumortissue (C) PCA of pituitary tumor tissue from an independent cohort using the 22 Func-PeLB probes showing that they segregate samples based on functional status in both tissue and serum. (D) Box plots of serum (l 383 eft) and tissue (right) mean methylation for each PT (functioning and nonfunctioning) and nontumors samples.

Considering the prognostic value reported in glioma or meningiomas, we surveyed serum-methylation markers specific to the invasion status of PT. Corroborating the findings reported in the tissue, we found slight serum differences between invasive and noninvasive groups (data not shown)^13,16,17,21^. However, the association of tissue- or serum-derived methylation groups with the criteria that better predict PT with higher risk to progress or recur remains to be elucidated ^13,18,38–43^.

The application of PeLB score is not intended to replace the standard approaches to diagnose and classify PT which, in most of the cases, is satisfactorily performed by clinical features, hormonal assessment in the blood/urine and on the imaging of the pituitary gland ^44^. However, these results provide evidence that serum cfDNA constitutes a reliable source of clinically relevant tumor-specific epigenetic signatures in PT as observed in other CNS tumors ^36^. Potentially, the specificity of PeLB probes could be helpful to distinguish PT from other rare primary or secondary sellar tumors whose diagnosis by morphologic and immunohistochemical approaches may be challenging, unavailable and/or inconclusive (e.g. craniopharyngioma variants, lymphoma, metastasis etc)^5,34,45,46^.

In conclusion, our results indicate that similar to malignant tumors, PT releases circulating tumor DNA that present specific methylation patterns, recapitulating molecular features detected in PT-tissue (e.g. adenohypophyseal lineage-related). Serum from patients with PT provides tumor-specific methylation signatures that allow the classification of samples into PT subtypes or non-PT groups. Finally, our preliminary results underpin the potential application of methylation profile in the serum-based liquid biopsy as a noninvasive approach to assess clinically relevant epigenetic features useful for clinical purposes in the management of patients (e.g. aggressiveness markers, actionable markers to guide future clinical trials to treat aggressive, resistant or recurrent PT etc).

## Methods

### Patients

– We conducted a retrospective analysis of a cohort comprised of archival serum and paired tissue (fresh-frozen) from 13 patients who underwent transsphenoidal surgery for the resection of invasive (n=5) or noninvasive (n=8) macroadenomas of different functional status and histological subtypes (9 nonfunctioning: 4 gonadotroph and 5 null cell and 4 functioning: 2 lactotroph, 1 corticotroph and 1 mixed GH/PRL/TSH) (Table 1).Criteria for invasiveness was based on Knosp grades 3–4 (n=4) or invasion of clivus (n=1)^47,48^. MRI assessment for size, and invasiveness classification was blindly and independently performed by two physicians from the Henry Ford Health System (HFHS)(TA, KPA). HFHS Pathologists provided a comprehensive pathology report on adenohypophyseal immunostaining, necrosis and quantification of markers of proliferation (Ki-67, mitotic counts, p53). Control serum was obtained from patients without PT (three epileptic patients and one with a nontumor condition). Control pituitary tissue was obtained from non-neoplastic pituitary harvested at autopsy (FFPE). We also generated serum methylome data from patients with glioma (n=114), meningiomas (n=6) and other CNS conditions (brain metastasis, 1 brain colloid cyst, 6 brain radiation necrosis) (Supplemental Table 2) The project was approved by the HFHS Institutional Review Board (IRB#10963) and patients consented to have their specimens used for research purposes. Publicly available methylome data from colorectal carcinoma was retrieved (CRC, n=2 pooled samples)^49^.

**Table 1:** Summary of the clinicopathological features from patients harboring pituitary tumors and nontumor conditions (serum)

| Feature | Nontumor (n=4) | NFPT (n=9) | FPT (n=4) |
| --- | --- | --- | --- |
| **Gender** |  |  |  |
| Female | 1 | 3 | 1 |
| Male | 2 | 6 | 3 |
| Unknown | 1 | 0 | 0 |
| **Age at surgery (years)** |  |  |  |
| Median (Lower and upper quartile) | 31 (22-60) | 65(57-68) | 52(46-57) |
| **2017 WHO/Hormone staining** |  |  |  |
| Gonadotroph/LH and/or FSH | - | 4 | - |
| Null Cell | - | 5 | - |
| Corticotroph/ACTH | - | - | 1 |
| Mixed/GH/PRL/TSH | - | - | 1 |
| Lactotroph/PRL | - | - | 1 |
| Densely lactotroph/PRL | - | - | 1 |
| **Invasion status** |  |  |  |
| Invasive | - | 5 | 2 |
| Noninvasive | - | 4 | 2 |
| **Size** |  |  |  |
| Micro_<1cm | - | 0 | 1 |
| Macro_≥1-4cm | - | 7 | 2 |
| Giant_≥4cm | - | 2 | 1 |

Legend - FPT: Functioning pituitary tumor. ACTH: Adrenocorticotrophic hormone; GH: Growth hormone; PRL: prolactin; LH: luteinizing hormone; FSH: follicle stimulating hormone

### Serum collection and processing

For the specimens originated from the HFHS, peripheral blood (15 mL) was drawn from each subject at the time of surgery before the tumor excision (transphenoidal).

Serum sample was separated within 1 hour from collection by centrifugation at 1,300 x g for 10 minutes at 20°C; aliquoted into up to five 2 mL cryovials and stored at –80°C until processing. The methods for the publicly available data is described in their respective manuscripts ^49^

### DNA isolation, quantification,and DNA methylation data generation

Tissue and serum DNA were extracted from 2.2–9.3mL aliquots of serum using the Quick-cfDNA Serum & Plasma Kit according to the manufacturer’s protocol (Zymo Research – catalog # D4076). DNA concentration was measured with Qubit (Thermo Fisher Scientific) /or with 4200 TapeStation (Agilent Technologies). The concentration of cfDNA in the serum was calculated by dividing the total amount of cfDNA extracted by the amount of serum used for extraction. We then converted the concentration of cfDNA in the serum (ng/mL) into haploid genome equivalents/mL by multiplying by a factor of 303 (assuming the mass of a haploid genome 3.3 pg) ^50^.

The extracted DNA (30–300 ng) was bisulfite-converted (Zymo EZ DNA methylation Kit; Zymo Research) and profiled using an Illumina Human EPIC array (HM850K), at the USC Epigenome Center, Keck School of Medicine, University of Southern California, Los Angeles, California. The raw DNA methylation data reported in this paper has been deposited to Mendeley Data at https://data.mendeley.com/datasets/cgrz6zztfg.

### DNA methylation pre-processing

Methylation array data was processed with the minfi package in R. The raw signal intensities were extracted from the *.IDAT files and corrected for background fluorescence intensities and red-green dye-bias using the function preprocess Noob as described by Triche et al., 2013 ^51^. The beta-values were calculated as (M/(M+U)), in which M and U refer to the (pre-processed) mean methylated and unmethylated probe signal intensities, respectively. Measurements in which the fluorescent intensity was not statistically significant above background signal (detection p value > 10^−16^) were removed from the data set. Before the analysis, we filtered out probes that were designed for sequences with known polymorphisms or probes with poor mapping quality (complete list of masked probes provided by Zhou et al.^52^) and the X and Y chromosomes.

### Deconvolution

We applied a previously described methodology ^50^ to deconvolute the relative contribution of cell types to a given sample ^50^. We included methylation signatures from cell lines, immune cells (B-cell, CD4T, CD8T, natural killer cells and white blood cells (monocytes, neutrophils) and vascular endothelial cells ^50^(Supplemental Table 2) For lack of information related to methylation signatures from individual cells that comprise the pituitary gland, we generated genome-wide methylation signatures from bulk non-neoplastic pituitaries obtained from cadavers (unpublished data) and followed the steps for defining the signatures as previously described ^50^. Briefly, we selected the 100 most specific hypermethylated and hypomethylated CpG probes for each cell/tissue type of interest. Using this signature, we applied a non-negative least squares method to deconvolute our serum and tissue cohort using the standalone program provided by Moss and colleagues ^50^. We then normalized the percentages generated by the standalone program for each cell type/PT-tissue from 0 to 100 by serum.

### DNA methylation exploratory analysis (unsupervised analysis)

In order to evaluate the DNA methylation profile in the serum from patients with distinct tumor types and non-neoplastic brain diseases, we performed a genome-wide Principal Component Analysis (PCA) across the samples (N=147) using the function *prcomp*(version 3.6.0). Consensus clustering was determined by k-means clustering of euclidean distance from the *ConsensusClusterPlus*(version 1.48.0) package.

### Supervised analysis

We also performed an epigenome-wide differential analysis across the serum from 10 patients with PT and 105 with non-PT conditions patients (4 non-tumor, 114 glioma, 3 meningioma, 1 brain metastasis carcinoma, 1 colloid cyst, and 4 from other CNS necrotic tumors). We used the Wilcoxon rank-sum test to identify differentially methylated probes between two different pairs: PT vs non-PT and functioning vs nonfunctioning PT.

For the comparison between PT and non-PT, probes were considered differentially methylated when the false discovery rate (FDR) was less than .001 and absolute value of the difference of a pair of probe mean methylation between each group was greater than 20%. To identify DMP in the serum that were tissue-specific, we calculated the differences in DNA methylation between the matching serum and tissue, by patient. We then selected probes with less than 5% difference between tissue and serum and considered them tissue-specific.

To validate their PT-specificity, we overlapped PeLB probes with the DNA methylome of an independent cohort consisting of pituitary-, glioma- and meningioma-tissue(Figure 1D).

For the comparison between functioning and nonfunctioning, probes were considered differentially methylated when the p-value was less than .01 and absolute value of the difference of probe mean methylation between each group was greater than 20%.

### Random Forest

We used a random forest machine-learning (ML) model for binary classification of the specimens with the aim to classify available cfDNA methylation (from serum) derived from patients with PT and non-PT (other neoplastic or non-neoplastic conditions: meningioma, glioma and colorectal carcinoma and nontumor). We first randomly allocated 20% of all samples for the validation set (n = 3 PT; n = 29 Non-PT) only analyzed for the assessment of the prediction model accuracy. The remainder serum specimens were used for the feature extraction or training of the random forest model. For developing the model we randomly partitioned the remainder samples into a training (n = 8 PT; n =84 Non-PT) and testing set (n = 2 PT; n = 21 Non-PT). We used the function *train* (package *caret* version 6.0.82) in CRAN, with 5000 trees, and 10 fold cross validation to generate our model. When testing the model, we used an output of 50% probability as a cut-off for classification.

Based on this result, we adopted the default PeLB score cutoff value of 50 to determine whether a patient had PT. We evaluated the performance of the prediction by applying the ML model on the validation set.

### Probe annotation

CpG probes were mapped to their CpG genomic location as CpG islands (CGI), shores, shelves, and open sea regions as previously defined ^52–55^.

### Statistical analysis

All processing and statistical analyses were done in R (3.6.1). Wilcoxon rank-sum test and multiple testing adjustments (e.g. FDR) were used to identify differentially methylated probes (DMP) as stated in the previous sections.

## Data Availability

The data will be available under the accession code GSEXXXX. All the other data supporting the findings of this study are available within the article and supplemental information and from the corresponding author upon reasonable request.

## Supplementary information

## Acknowledgements

The authors are grateful to the HFHS patients who consented to the usage of PT for research purposes. We thank Nancy Takacs and Heather Mengel for their administrative support; Kevin Nelson for the collection, handling and maintenance of the tumor bank at the Hermelin Brain Tumor Center; Andrea Transou for tumor pathology processing; Laura A. Hasselbach for DNA extraction; Daniel Weisenberger and team at USC Epigenome Center for assistance with DNA methylation profiling (HFHS support);Susan MacPhee for proofreading the manuscript.

## Author contributions

Overall concept and coordination of the study: AVC, JR, HN, KPA; retrieval of publicly available molecular and clinical data: KPA, MW, AVC; Bioinformatic and statistical analyses: MW, TSS, TMM, MSM, HN and input from LMP; HFHF cohort: pathology review AM, DC; molecular data generation: TMM, AD; the manuscript was written by AVC, HN, MW and intellectual contribution from JS, TM, SK, TW. All authors contributed to the revision of the manuscript.

## Data availability

The data is available under the accession code GSEXXXX. All the other data supporting the findings of this study are available within the article and supplemental information and from the corresponding author upon reasonable request.

## Footnotes

These authors contributed equally as first authors: Michael Wells, Karam P. Asmaro,Thais S. Sabedot, Tathiane M. Malta, and Maritza S. Mosella. These authors contributed equally as senior authors: Houtan Noushmehr, Ana Valeria Castro

## Contributor information

Ana Valeria Castro

Houtan Noushmehr, contributed to the revision of the manuscript. the findings of this study are available within the article and supplemental information and from the corresponding author upon reasonable request.

## SUPPLEMENTAL INFORMATION

**Table s1**:–(A)Distribution of the number of the most variant methylated probes in the serum and in the pituitary tumor tissue according to their genomic context and CpG location. (B) Distribution of the number of differentially methylated probes(PeLB and Func-PeLB) according to their genomic context and CpG location.

**Figure s1** –(A)Cell-free DNA concentration in the serum derived from patients with nontumor and tumor conditions. (B) Cell-free DNA concentration in the serum of patients with pituitary tumors (PT) according to function and invasion status.(C)Deconvolution of serum in relation to cell composition from control or pituitary tumor patients.(D) Workflow displaying the selection criteria of pituitary tumor- and functioning-specific methylated probes from the supervised analysis between PT and non-PT (right) and Functioning and nonfunctioning PT (left) in the serum. (E) Heatmap displaying the genome-wide methylation profile across PT tissue-specimens.

## Bibliography

1. Megnis, K. et al. Evaluation of the possibility to detect circulating tumor DNA from pituitary adenoma. Front Endocrinol (Lausanne) 10, 615 (2019).

2. Mouliere, F. et al. High fragmentation characterizes tumour-derived circulating DNA. PLoS ONE 6, e23418 (2011).

3. Jiang, P.& Lo, Y. M. D. The Long and Short of Circulating Cell-Free DNA and the Ins and Outs of Molecular Diagnostics. Trends Genet. 32, 360–371(2016).

4. Heitzer, E.& Speicher, M. R. One size does not fit all: Size-based plasma DNA diagnostics. Sci. Transl. Med. 10,(2018).

5. Shen, S. Y. et al. Sensitive tumour detection and classification using plasma cell-free DNA methylomes. Nature 563, 579–583(2018).

6. Kim, H., Wang, X.& Jin, P. Developing DNA methylation-based diagnostic biomarkers. J. Genet. Genomics 45, 87–97(2018).

7. Wang, J.& Bettegowda, C. Applications of DNA-Based Liquid Biopsy for Central Nervous System Neoplasms. J. Mol. Diagn. 19, 24–34(2017).

8. Best, M. G. et al. Liquid biopsies in patients with diffuse glioma. Acta Neuropathol. 129, 849–865(2015).

9. Adamczyk, L. A. et al. Current Understanding of Circulating Tumor Cells – Potential Value in Malignancies of the Central Nervous System. Front. Neurol. 6, 174 (2015).

10. Saenz-Antoñanzas, A. et al. Liquid biopsy in glioblastoma: opportunities, applications and challenges. Cancers (Basel) 11,(2019).

11. Bos, M. K. et al. Whole exome sequencing of cell-free DNA – A systematic review and Bayesian individual patient data meta-analysis. Cancer Treat. Rev. 83, 101951 (2020).

12. Koeppel, F. et al. Whole exome sequencing for determination of tumor mutation load in liquid biopsy from advanced cancer patients. PLoS ONE 12, e0188174 (2017).

13. Neou, M. et al. Pangenomic classification of pituitary neuroendocrine tumors. Cancer Cell(2019). doi:10.1016/j.ccell.2019.11.002

14. Bi, W. L. et al. Landscape of genomic alterations in pituitary adenomas. Clin. Cancer Res. 23, 1841–1851(2017).

15. Song, Z.-J. et al. The genome-wide mutational landscape of pituitary adenomas. Cell Res. 26, 1255–1259(2016).

16. Ling, C. et al. A pilot genome-scale profiling of DNA methylation in sporadic pituitary macroadenomas: association with tumor invasion and histopathological subtype. PLoS ONE 9, e96178 (2014).

17. Kober, P. et al. DNA methylation profiling in nonfunctioning pituitary adenomas. Mol. Cell. Endocrinol. 473, 194–204(2018).

18. Salomon, M. P. et al. The Epigenomic Landscape of Pituitary Adenomas Reveals Specific Alterations and Differentiates Among Acromegaly, Cushing’s Disease and Endocrine-Inactive Subtypes. Clin. Cancer Res. 24, 4126–4136(2018).

19. Duong, C. V. et al. Quantitative, genome-wide analysis of the DNA methylome in sporadic pituitary adenomas. Endocr. Relat. Cancer 19, 805–816(2012).

20. Capper, D. et al. DNA methylation-based classification of central nervous system tumours. Nature 555, 469–474(2018).

21. Gu, Y.et al. Differential DNA methylome profiling of nonfunctioning pituitary adenomas suggesting tumour invasion is correlated with cell adhesion. J Neurooncol 129, 23–31(2016).

22. Zhou, Y., Zhang, X.& Klibanski, A. Genetic and epigenetic mutations of tumor suppressive genes in sporadic pituitary adenoma. Mol. Cell. Endocrinol. 386, 16–33(2014).

23. Torregrosa-Quesada, M. E. et al. How Valuable Is the RT-qPCR of Pituitary-Specific Transcription Factors for Identifying Pituitary Neuroendocrine Tumor Subtypes According to the New WHO 2017 Criteria? Cancers (Basel) 11,(2019).

24. Moran, S. et al. Epigenetic profiling to classify cancer of unknown primary: a multicentre, retrospective analysis. Lancet Oncol. 17, 1386–1395(2016).

25. Hoadley, K. A. et al. Cell-of-Origin Patterns Dominate the Molecular Classification of 10,000 Tumors from 33 Types of Cancer. Cell 173, 291–304.e6(2018).

26. Ceccarelli, M. et al. Molecular profiling reveals biologically discrete subsets and pathways of progression in diffuse glioma. Cell 164, 550–563(2016).

27. Sahm, F. et al. DNA methylation-based classification and grading system for meningioma: a multicentre, retrospective analysis. Lancet Oncol. 18, 682–694(2017).

28. Ferraresso, S. et al. DNA methylation profiling reveals common signatures of tumorigenesis and defines epigenetic prognostic subtypes of canine Diffuse Large B-cell Lymphoma. Sci. Rep. 7, 11591 (2017).

29. Mosella, M. S. et al. DNA Methylation-based Signatures Classify Sporadic Pituitary Tumors According to Clinicopathological Features. BioRxiv (2020). doi:10.1101/2020.04.25.061903

30. Lokk, K. et al. DNA methylome profiling of human tissues identifies global and tissue-specific methylation patterns. Genome Biol. 15, r54 (2014).

31. Yang, X., Gao, L.& Zhang, S. Comparative pan-cancer DNA methylation analysis reveals cancer common and specific patterns. Brief. Bioinformatics 18, 761–773(2017).

32. Hao, X. et al. DNA methylation markers for diagnosis and prognosis of common cancers. Proc Natl Acad Sci USA 114, 7414–7419(2017).

33. Widschwendter, M. et al. Methylation patterns in serum DNA for early identification of disseminated breast cancer. Genome Med. 9, 115 (2017).

34. Constâncio, V. et al. Early detection of the major male cancer types in blood-based liquid biopsies using a DNA methylation panel. Clin. Epigenetics 11, 175 (2019).

35. Kang, S. et al. CancerLocator: non-invasive cancer diagnosis and tissue-of-origin prediction using methylation profiles of cell-free DNA. Genome Biol. 18, 53 (2017).

36. Noushmehr, H. et al. Detection of glioma and prognostic subtypes by non-invasive circulating cell-free DNA methylation markers. BioRxiv(2019).doi:10.1101/601245

37. Mete, O.& Lopes, M. B. Overview of the 2017 WHO classification of pituitary tumors. Endocr. Pathol. 28, 228–243(2017).

38. Raverot, G. et al. Risk of Recurrence in Pituitary Neuroendocrine Tumors: A Prospective Study Using a Five-Tiered Classification. J. Clin. Endocrinol. Metab. 102, 3368–3374(2017).

39. Fernández-Balsells, M. M. et al. Natural history of nonfunctioning pituitary adenomas and incidentalomas: a systematic review and metaanalysis. J. Clin. Endocrinol. Metab. 96, 905–912(2011).

40. Vasiljevic, A., Jouanneau, E., Trouillas, J.& Raverot, G. Clinicopathological prognostic and theranostic markers in pituitary tumors. Minerva Endocrinol. 41, 377–389(2016).

41. Selman, W. R., Laws, E. R., Scheithauer, B. W.& Carpenter, S. M. The occurrence of dural invasion in pituitary adenomas. J. Neurosurg. 64, 402–407(1986).

42. Asioli, S. et al. Validation of a clinicopathological score for the prediction of post-surgical evolution of pituitary adenoma: retrospective analysis on 566 patients from a tertiary care centre. Eur. J. Endocrinol. 180, 127–134(2019).

43. Trouillas, J. et al. A new prognostic clinicopathological classification of pituitary adenomas: a multicentric case-control study of 410 patients with 8 years post operative follow-up. Acta Neuropathol. 126, 123–135(2013).

44. Melmed, S. Pituitary-Tumor Endocrinopathies. N. Engl. J. Med. 382, 937–950(2020).

45. Nunes, S. P. et al. Subtyping lung cancer using DNA methylation in liquid biopsies. J. Clin. Med. 8,(2019).

46. Roy, D.& Tiirikainen, M. Diagnostic power of DNA methylation classifiers for early detection of cancer. Trends Cancer 6, 78–81(2020).

47. Micko, A. S. G., Wöhrer, A., Wolfsberger, S.& Knosp, E. Invasion of the cavernous sinus space in pituitary adenomas: endoscopic verification and its correlation with an MRI-based classification. J. Neurosurg. 122, 803–811(2015).

48. Knosp, E., Steiner, E., Kitz, K.& Matula, C. Pituitary adenomas with invasion of the cavernous sinus space: a magnetic resonance imaging classification compared with surgical findings. Neurosurgery 33, 610–7; discussion 617 (1993).

49. Gallardo-Gómez, M. et al. A new approach to epigenome-wide discovery of non-invasive methylation biomarkers for colorectal cancer screening in circulating cell-free DNA using pooled samples. Clin. Epigenetics 10, 53 (2018).

50. Moss, J. et al. Comprehensive human cell-type methylation atlas reveals origins of circulating cell-free DNA in health and disease. Nat. Commun. 9, 5068 (2018).

51. Triche, T. J., Weisenberger, D. J., Van DenBerg, D., Laird, P. W.& Siegmund, K. D. Low-level processing of Illumina Infinium DNA Methylation BeadArrays. Nucleic Acids Res. 41, e90 (2013).

52. Zhou, W., Laird, P. W.& Shen, H. Comprehensive characterization, annotation and innovative use of Infinium DNA methylation BeadChip probes. Nucleic Acids Res. 45, e22 (2017).

53. Gardiner-Garden, M.& Frommer, M. CpG islands in vertebrate genomes. J. Mol. Biol. 196, 261–282(1987).

54. Sandoval, J. et al. Validation of a DNA methylation microarray for 450,000 CpG sites in the human genome. Epigenetics 6, 692–702(2011).

55. Takai, D.& Jones, P. A. The CpG island searcher: a new WWW resource. In Silico Biol (Gedrukt) 3, 235–240(2003).

